# Using wastewater surveillance to explore community-level dietary intake in sewered and non-sewered sanitation systems in Malawi, Africa

**DOI:** 10.64898/2026.06.07.26354900

**Authors:** Rochelle H. Holm, Petros Chigwechokha, Chisomo Kaponda, Makayla Stephens, Hannah Limbong, Ayse Ercumen, Joy L. Hart, Cassandra L. Workman, Francis L. de los Reyes, Brighton A. Chunga

## Abstract

Wastewater can be used to measure biomarkers that reflect population-level dietary intake and diversity; however, how this approach may apply in a low-income country remains a knowledge gap. This study aims to evaluate whether select dietary-related metabolites can be detected in wastewater and environmental surveillance (WES) samples from both sewered and non-sewered sanitation systems in Malawi, Africa. Fourteen WES samples were collected and analyzed from two university campuses in Mzuzu and Thyolo, Malawi. Four targets were analyzed: N-methyl-2-pyridone-5-carboxamide (2PY; a biomarker of vitamin B3), 4-pyridoxic acid (4-PA; a biomarker of vitamin B6), as well as enterodiol and enterolactone (biomarkers of dietary fiber and polyphenol consumption). An 18-question survey, paired spatiotemporally with the WES measurements, assessed self-reported daily dietary intake, food insecurity, and nutrient deficiency symptoms among 500 respondents. Among the 14 WES samples, 2PY, 4-PA, and enterolactone were detected, while enterodiol was not detected above the method limit (<0.3 mg/kg). Most respondents (79%; 397/500) reported consuming foods associated with the 2PY biomarker. Many respondents (62%; 311/500) also reported consuming foods linked to the 4-PA biomarker. Fewer respondents (36%; 181/500) reported consuming foods associated with enterodiol or enterolactone, such as whole grains (e.g., millet) and other fiber-rich plant foods (e.g., beans, chickpeas, or pigeon peas). This study demonstrates the potential feasibility of monitoring dietary-related metabolites in both sewered and non-sewered sanitation systems in a low-income country to augment community-level nutrition data. 2PY, 4-PA, and enterolactone were detectable in WES samples, supporting the advancement of this emerging field in nutrition and food security research.

## Introduction

Wastewater and environmental surveillance (WES) can yield valuable health data for understanding diseases, diet, and environmental exposures within communities.^1–4^ WES samples can be collected from both sewered and non-sewered sanitation systems and can provide insights into various public health parameters, which can then be used to inform community-level therapeutic decisions together with, or in the absence of, available clinical data.^5^ Traditional methods for tracking diet, such as questionnaires, food diaries, and stool samples, are often resource-intensive and collected at an individual level; these methods are limited in providing a rapid assessment of a population’s health status. Building on approaches that detect urine and stool biomarkers indicating dietary intake in high-income countries,^1,3^ WES in low- and middle-income countries (LMICs), which may be more food insecure, offers the potential to support community-level nutrition status evaluations.

Specifically, wastewater can be used to measure population-level dietary biomarkers of vitamins B3 (niacin)^3,6^ and B6 (pyridoxine),^3,6,7^ as well as fiber intake from whole grains and other fiber-containing plant foods using enterodiol^3,6,8^ or enterolactone.^3,6^ N-methyl-2-pyridone-5-carboxamide (2PY) is a urinary biomarker of vitamin B3 over the past 2–4 days.^9^ Vitamin B3 food sources include bran, yeast, eggs, peanuts, poultry, red meat, fish, whole-grain cereals, legumes, and seeds; low vitamin B3 intake is associated with pellagra and symptoms of diarrhea, dermatitis, dementia, and death.^10^ 4-pyridoxic acid (4-PA) is a direct vitamin B6 biomarker over the past 2–4 days, with about 40–60% of dietary vitamin B6 excreted in urine as 4-PA.^9,11^ Vitamin B6 food sources are typically protein-rich foods (e.g., meat, liver, fish, beans, and lentils). Low vitamin B6 intake in the general population is associated with a higher risk of cardiovascular disease and cancer.^11^ Enterolactone has shown high interindividual variability, but urinary excretion at 48 h post-meal results in a high recovery of the plant lignans consumed.^12^ The urinary excretion concentration of enterolactone tends to be higher than that of enterodiol.^13^ 2PY, 4-PA, enterodiol, and enterolactone have been shown to be detectable in wastewater and are moderately stable for WES applications.^14,15^

Public opinion in Malawi, Africa, has supported WES as a potential tool for dietary surveillance.^16^ The Malawian diet, which is generally carbohydrate-dense, relies heavily on smallholder maize farming, leaving the population susceptible to malnutrition due to subsistence production and lack of nutrient balance. Access to imported food products is limited, and imported food is generally unaffordable for most people; locally grown foods therefore comprise the majority of intake. Malnutrition is particularly severe among children under five, with 38% stunted, 10% underweight, and 2% wasted.^17^ Tembo et al.^18^ found that culture and religion largely dictate dietary preferences in Malawi. Additionally, food insecurity among tertiary education students is a worldwide problem; after paying for books, tuition, and housing (plus other essential expenses), some students lack the money to buy food.^19^

This study aims to determine whether select dietary-related metabolites can be detected in WES samples from both sewered and non-sewered sanitation systems at two university campuses in Malawi. Four targets were analyzed: N-methyl-2-pyridone-5-carboxamide (2PY), 4-pyridoxic acid (4-PA), enterodiol, and enterolactone. Findings from this work can inform whether WES-based dietary-related metabolites can be extended to LMICs.

## Materials and Methods

### Study sites

The study was conducted on the campuses of two public universities: Mzuzu University (MZUNI) in the northern region and Malawi University of Science and Technology (MUST) in the southern region. These two sites were selected to represent distinct locations across Malawi and to capture many adults living in congregate settings. MZUNI is located in Mzuzu and has approximately 10,000 students living both on campus and in the surrounding community. MUST is located in Thyolo and has approximately 8,000 students, most of whom reside on campus. Wastewater samples and concurrent dietary survey data were collected when classes were in session (May–June 2025). This study period coincided with the maize harvest season, when food is generally more available.^20,21^ The diet of Malawi residents shows limited regional differences, and sampling during the food-secure season increased the likelihood of detecting the wastewater targets.

### Recruitment

Food intake and food security data were collected in person using paper-based surveys. The survey was piloted with approximately 10 respondents prior to data collection in the Mzuzu area. The target sample size was 500 participants, with 250 respondents at each study site. Participants were purposively recruited on campus to include both staff and students. A geofencing approach ensured that surveys were conducted only within campus boundaries, aligning with the wastewater sampling areas. Respondents were recruited through convenience sampling in academic buildings, near student accommodations, and in outdoor leisure spaces. Eligibility was limited to adults aged 18 years or older. Surveys were administered in English or Chichewa, the two official languages of Malawi.

### Dietary survey

The 18-question survey (Supplemental Material Text S1) aimed to assess daily dietary intake of food categorized into eight groups for correlation with the four targets, food insecurity of the respondent and their household, and self-reported nutrient deficiency symptoms potentially linked to the four wastewater targets. Survey responses included multiple-choice, open-ended, Likert-scale, and yes/no formats. Self-reported symptoms linked to deficiencies associated with the four wastewater targets under study included physical and cognitive symptoms over the last 4 weeks, such as fatigue or low energy, muscle weakness, and headache, and were ranked on a Likert scale from Never (0) to Often (4). Respondents also provided demographic information, including age group, gender, education level, and the type of toileting facility most frequently used. Questions were either read aloud by study personnel or completed independently by participants. The average completion time ranged from 15–20 min. No incentives were offered to participants.

### Wastewater sample collection

Grab samples were collected once per day at each site for seven consecutive days at both MZUNI and MUST. At each sampling point, 2 L of wastewater were collected, transported on ice packs to the laboratory, and stored at 4 °C. Samples were processed by hand shaking to homogenize them, after which 50 mL aliquots were transferred into centrifuge tubes and centrifuged for 15 min. The supernatant was discarded. This process was repeated until a total of 15 g of solid material was obtained for each sampling day. Samples were stored at 4 °C until analysis.

At MZUNI, samples were collected from an open drain that primarily received effluent from residential student housing serving several hundred students. The sampling site was <100 m from student housing. The MZUNI campus includes a mix of sewered and non-sewered systems (septic tanks and pit latrines). At MUST, samples were collected at a pump station that aggregated wastewater from student housing (∼8,000 students) and the wider campus prior to treatment. This campus is served by a piped system leading to an onsite treatment pond. The sampling site was 30–<300 m from the student housing buildings.

During the study period, 24 h composite sampling was not feasible at either site due to the remoteness of the locations. Based on discussions with university facilities personnel, no industrial wastewater was reported upstream, and animal input was assumed to be minimal. Flow data were unavailable, but both locations had continuous liquid flow throughout the sampling period.

### Wastewater analysis

Wastewater pellets were analyzed at the Malawi Bureau of Standards for four target nutrition biomarker indicators: 2PY, 4-PA, enterodiol, and enterolactone (Table 1). We chose just four biomarkers to represent a wide range of food sources that varies greatly across each food group or indicator and is not indicative of all of those food sources.

**Table 1.** Targets studied in wastewater and environmental surveillance, sources in local Malawian foods, and associated deficiency symptoms.

| Target | Biomarker for | Source in local food | Deficiency symptoms |
| --- | --- | --- | --- |
| N-methyl-2-pyridone-5-carboxamide (2PY) | Vitamin B3 (niacin) | bran, yeast, eggs, peanuts, poultry, red meat, fish, whole-grain cereals, legumes, and seeds | Pellagra (diarrhea, dermatitis, dementia and death) |
| 4-pyridoxic acid (4-PA) | Vitamin B6 (pyridoxine) | meat, liver, fish, beans, and lentils | higher risk of cardiovascular disease and cancer |
| Enterodiol | Fiber & healthy diet | whole grains and other fiber-containing plant foods | constipation, weight gain, unstable blood sugar levels, and bloating |
| Enterolactone | Fiber & healthy diet | whole grains and other fiber-containing plant foods | constipation, weight gain, unstable blood sugar levels, and bloating |

A UHPLC KNAUER system with a diode array detector (DAD) was used for the analysis. A 50 mL Falcon tube was filled with 2 g of wastewater pellet material. In brief, samples for 2PY and 4-PA were prepared based on a modified Food & Beverage Testing method.^22^ Standards were prepared in HPLC-grade water. The wavelength for detection was 296 nm; the isocratic mobile phase was water:acetonitrile (90:10 v/v); temperature was 40 °C; and the flow rate was 0.7 mL/min. For enterodiol and enterolactone, samples were prepared based on a modified Food & Beverage Testing method,^22^ but water was substituted with HPLC-grade methanol for both samples and standards. The wavelength for detection was 280 nm; temperature was 40 °C; the flow rate was 1.0 mL/min; mobile phase A was water:methanol (60:40 v/v); and mobile phase B was methanol:water (70:30 v/v).

The following gradient program was used: 1.0–6.9 min, A = 100%; 7.1–7.5 min, B = 100%; 7.6–10 min, A = 100%. The calibration curve consisted of seven levels. The retention time for 4-PA was 1.7 min; 2PY was 2.0 min; enterolactone was 7.45 min; and enterodiol was 8.45 min.

### Data analysis

The survey results were manually entered into Excel and verified by two researchers. Descriptive analysis of dietary intake and nutrient deficiency symptoms was conducted. Differences in wastewater concentrations for the four dietary-related metabolites between the two study sites were assessed using a Mann–Whitney U test; the primary study hypothesis was that the two sites would not differ from each other. A linear correlation analysis was also conducted. Analyses were conducted using R version 4.5.2 and RStudio (2026.01.0+392).^23,24^

### Ethics

This study was reviewed and approved by the Malawi University of Science and Technology Research Ethics Committee (P.01/2024/117; 17 April 2025) and the University of Louisville Institutional Review Board (25.0281). Written informed consent was obtained from all survey participants.

## Results and Discussion

Complete survey responses were obtained from 500 participants (S1 File: Dietary survey full results). The respondents were predominantly male (63%; 316/500) and younger than 34 years (91%; 457/500). About half (53%; 267/500) had completed secondary school, and nearly equal numbers reported using a flush toilet (47%; 233/500) or a pit latrine (47%; 237/500) as their primary toileting facilities. Among the 14 WES samples analyzed for the four targets, three were detected: 2PY (mean, 72.9 mg/kg; 95% CI: 54.8–91.0 mg/kg), 4-PA (mean, 5.0 mg/kg; 95% CI: 3.2–6.7 mg/kg), and enterolactone (mean, 6.7 mg/kg; 95% CI: 4.9–8.6 mg/kg). Enterodiol was not detected above method limits (<0.3 mg/kg) (Figure 1). Of the four biomarkers, enterolactone showed a wider concentration range in wastewater, consistent with its high individual variability^12^. There was no correlation between the two vitamin B targets (4-PA and 2PY; R² = 0.0015). Similarly, there was no relationship between enterolactone and either of the vitamin B targets (4-PA and enterolactone, R² = 0.0016; 2PY and enterolactone, R² = 0.0813).

**Figure 1.**
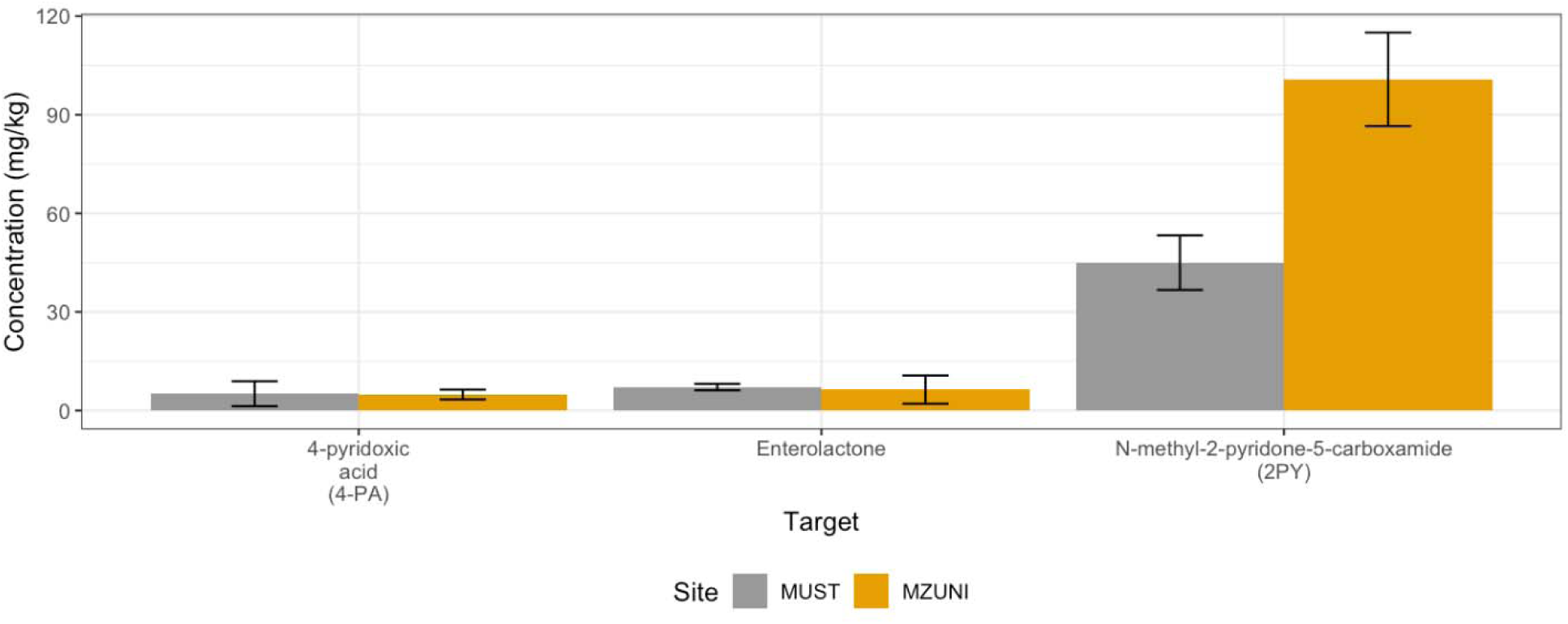
Summary concentrations of N-methyl-2-pyridone-5-carboxamide (2PY), 4-pyridoxic acid (4-PA), and enterolactone in wastewater and environmental samples from the two study sites (Mzuzu University and Malawi University of Science and Technology) (N = 14). Error bars represent 95% confidence intervals.

The stability of these biomarkers in wastewater is dependent on the hydraulic detention time, the time from the generation site to the sampling point.^14,15^ The four biomarkers in this study were moderately stable, having reported half-lives of 3.8 h for 2PY, 7.8 h for 4-PA, 8.6 h for enterodiol, and 6.9 h for enterolactone in piped sewer systems.^15^ However, the stability in a non-sewered system, such as the MZUNI site, has not been established. When comparing the two study sites (MZUNI and MUST), wastewater concentrations were similar for 4-PA (p-value = 0.80) and enterolactone (p-value = 0.34). However, significant differences were observed for 2PY between the two sites (p-value < 0.001), which may be due to sampling an open drain versus a piped system. This is also likely due to the hydraulic detention time being shorter at MZUNI and the lower half-life of 2PY compared with 4-PA and enterolactone. It is also possible that there is greater degradation in a piped system. Additionally, although reported to be moderately stable in wastewater,^15^ enterodiol is an intermediate human gut microbial transformation that is eventually converted to enterolactone in most cases; thus, it makes sense that enterodiol was not detected above the method limits.

Food variety is an important consideration in food security, with food-insecure people tending to eat a narrower range of foods.^25,26^ Of the 107 total individual foods across the eight food categories, MUST respondents reported eating an average of 14 different foods, and MZUNI respondents reported eating an average of 11 different foods. Most respondents (78%; 388/500) reported maize as their main household food source, consistent with previous diet surveys in the study area.^20,21^ Additionally, 79% (397/500) reported consuming one or more foods associated with the 2PY biomarker, such as bran, yeast, eggs, peanuts, poultry, red meat, fish, whole-grain cereals (e.g., millet), legumes, and seeds. This category (Table 1) represented the broadest range of foods in the study and corresponded to the widest range of wastewater concentrations. Many respondents (62%; 311/500) also consumed one or more foods linked to the 4-PA biomarker, including chicken, beef, liver, fish, beans, and lentils. Fewer respondents (36%; 182/500) reported consuming one or more foods associated with enterodiol or enterolactone, such as whole grains (e.g., millet) and other fiber-rich plant foods (e.g., beans, chickpeas, or pigeon peas). In addition to being a food consumption biomarker, 4-PA has also been used as a population size normalization biomarker.^7^

The survey results on vitamin deficiency experienced over the past four weeks showed that 13% (65/500) of the participants reported experiencing one or more symptoms always (Likert scale 4), and a total of 29% of participants experienced symptoms either always or often over the past four weeks (146/500). There was no difference between MUST and MZUNI respondents reporting symptoms either always or often (p-value = 0.96). The MZUNI wastewater sample included a smaller population size and might have been expected to have lower levels of biomarkers than MUST samples. However, we found the opposite for 2PY. Because biomarker concentrations could not be normalized to flow rates as a proxy for the number of people contributing to the wastewater samples, it is difficult to assess the correlation between 2PY and the nutritional status of the community. The non-random sampling approach means that these differences could have been affected by the hundreds of adequately nourished students at each campus. In many settings, traditional nutrition testing and evaluation methods have limited reach due to their resource-intensive nature, underreporting, and delays in data collection, making them less effective for early detection and response to emerging health threats. Regional community monitoring through WES, rather than requiring malnourished individuals to visit healthcare facilities, could support improved nutrition and health outcomes in LMICs if normalized to the population.

There is some evidence of UHPLC-MS/MS quantitative analytical method validation issues with enterodiol in environmental samples.^8^ Whereas enterolactone was detected in all WES samples in this study, Qi et al.^8^ similarly reported a high frequency of detection, particularly associated with mammalian urine and fecal excretions from leaking septic systems and discharge from a water treatment plant. The traditional food nsima, made from maize flour, contains vitamin B3 (niacin). This makes wastewater analysis of 2PY more challenging in the Malawi context, as almost the entire population has access to and consumes maize. In addition, the effects of population factors such as diet or diseases (e.g., kidney diseases) on wastewater concentration levels are not well understood.

This study is the first to examine the feasibility of monitoring dietary-related metabolites in both sewered and non-sewered sanitation systems in an LMIC. The availability and accessibility of food in the urban environment significantly influence health outcomes in LMICs, and the accessibility of food in one’s neighborhood is particularly important for diet.^27^ WES can potentially be used to identify malnutrition hotspots and areas for targeted interventions to encourage demand for nutritious diets. Because the diets in the two areas are nearly identical, the differences in the biomarker may reflect a broader dynamic range.

Using WES for surveillance of healthy eating and the incidence of malnutrition is in its infancy but can potentially be used alongside other community health indicators, such as pathogens (e.g., *Vibrio cholerae*, SARS-CoV-2). The same WES sample may be analyzed using multiple methods on a weekly or monthly schedule, providing a more complete picture of community health year-round.

### Limitations

The two study sites were institutions of higher education, and some staff members who were not students but may have used the toilet facilities were included. Access to secondary and tertiary education in Malawi involves significant barriers, and the student population does not fully represent the broader Malawian population. The non-detection of enterodiol may reflect a combination of dietary intake patterns, analytical sensitivity limits, degradation within wastewater matrices, or matrix suppression effects, and thus should be interpreted cautiously. To increase detection and demonstrate feasibility, solids were used for analysis, and the liquid fraction was discarded. This is a known issue, as 2PY and 4-PA are water-soluble, and the amount lost in the discarded supernatant is unknown. Better dietary recall focusing on dietary diversity and consumption volume would also help better characterize the food situation at the sites. Due to the nature of WBE in a low-resource LMIC, we were only able to collect grab samples and flow was not measured.

### Future research

This proof-of-principle study highlights several future global research needs: 1) The significant difference in 2PY levels at the two sample sites points to the need to investigate the effects of factors such as regional dietary differences, system differences between an open drain and piped sampling points, and hydraulic detention time; 2) Sampling is needed over time across a broader range of communities to establish baseline wastewater data, delineate the “normal” dynamic range of wastewater concentrations, and determine threshold levels of concern (e.g., seasonal variation in the biomarkers could be evaluated against the food availability during those seasons); 3) Comparison of wastewater samples from different regions with and without known food insecurity (e.g., a refugee camp or drought-ravaged area vs. a more affluent neighborhood), as well as comparison of sewered and non-sewered sampling points, would be insightful; 4) Metagenomic approaches for estimating dietary intake from stool samples, which are more effective for inferring whole-food intake,^1,28^ could also be used. Further investigation is needed to assess how these stool-based methods translate to African diets and wastewater matrices; and 5) Additional food consumption biomarkers with evidence of stability should be considered,^15^ including 3-carboxy-4-methyl-5-propyl-2-furanpropionic acid (CMPF), derived from meat-based foods, and gamma-carboxyethyl hydrochroman (γ-CEHC), associated with vitamin E intake.

## Conclusion

WES has the potential to provide a community-level snapshot of population dietary-related diversity. This proof-of-concept study demonstrates the feasibility of wastewater surveillance as a novel approach for tracking dietary-related metabolites in an LMIC with both in sewered and non-sewered sanitation systems. 2PY, 4-PA, and enterolactone were detectable in WES samples, supporting the advancement of this emerging field in nutrition research.

## Availability of data and materials

All data generated or analyzed during this study are included in this article and its supplementary information files.

## Authors’ contributions

R.H.H. and P.C. conceived and designed the research; C.K., M.S. and H.L. collected field samples; R.H.H., P.C. and C.K. analyzed data; R.H.H. drafted the manuscript; R.H.H., P.C., C.K., M.S., H.L., A.E., J.L.H., C.L.W., F.L.D., and B.A.C. edited and revised manuscript; R.H.H., P.C., C.K., M.S., H.L., A.E., J.L.H., C.L.W., F.L.D., and B.A.C. approved final version of manuscript.

## Supplementary Materials

**Figure S1.**
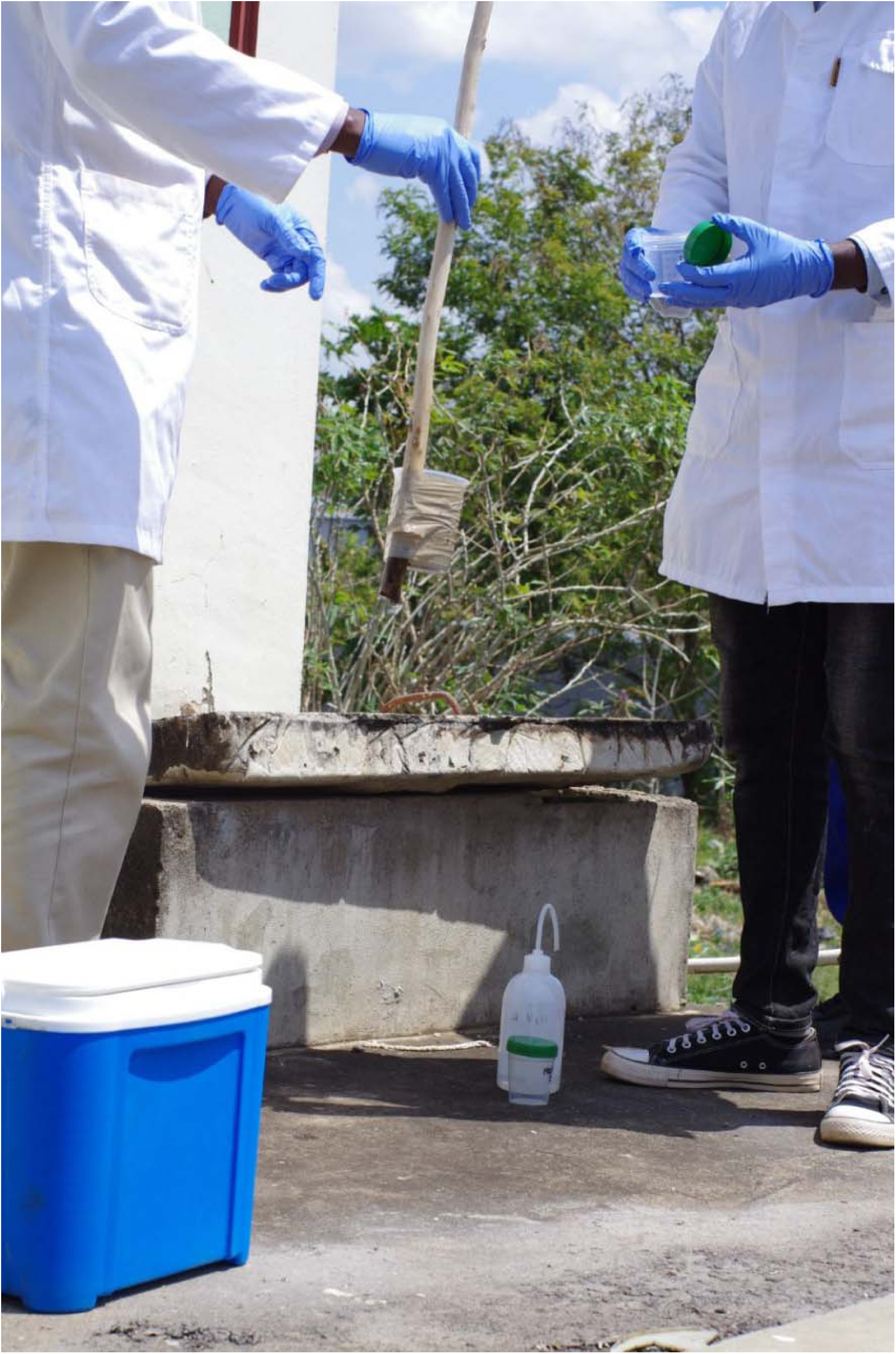
Grab sample collection at Malawi University of Science and Technology. The sampling site was a pump station that aggregated wastewater from student housing and the wider campus prior to treatment.

**Figure S2.**
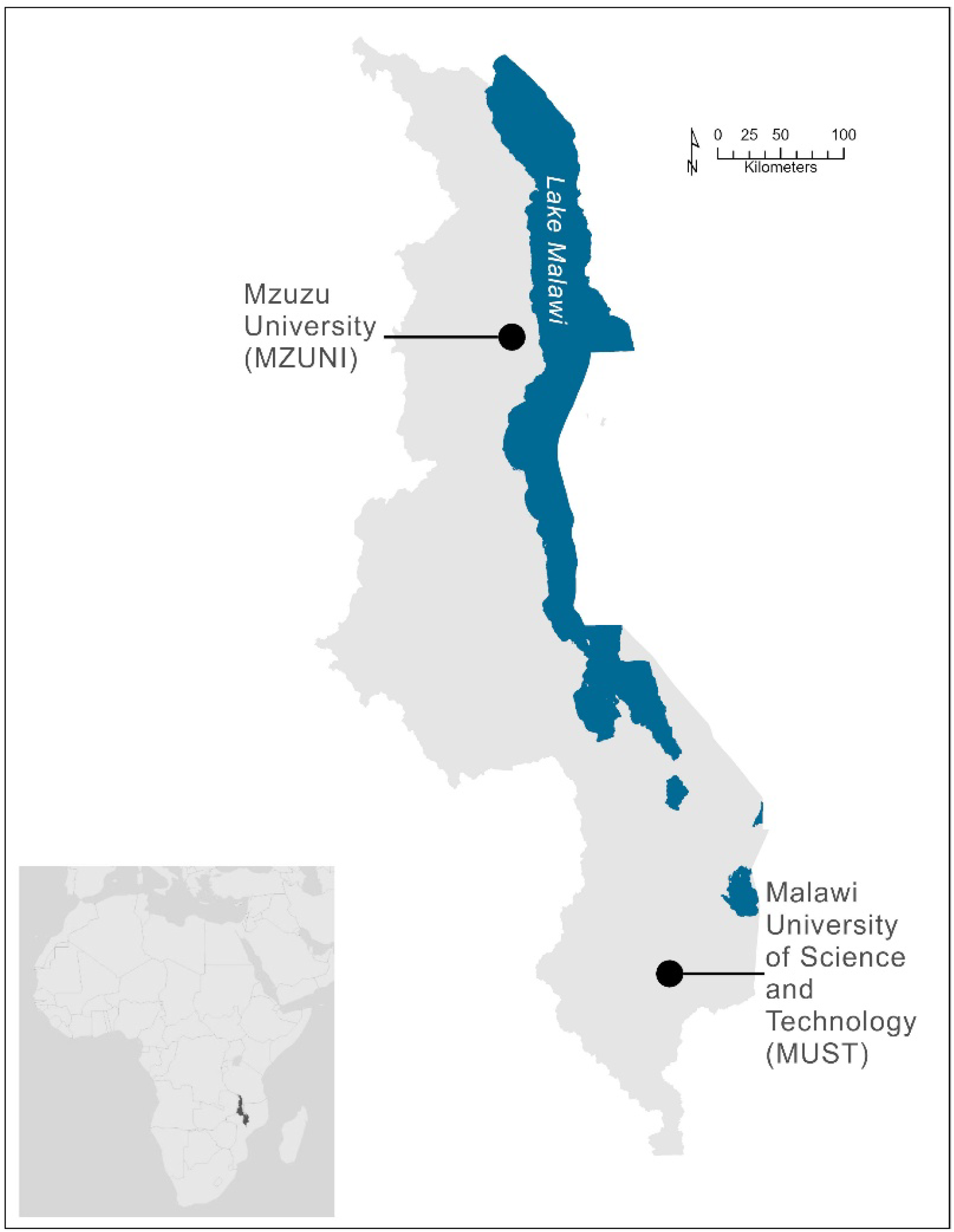
Two study sites: Mzuzu University and Malawi University of Science and Technology, Malawi, Africa.

---

**Table S1.**
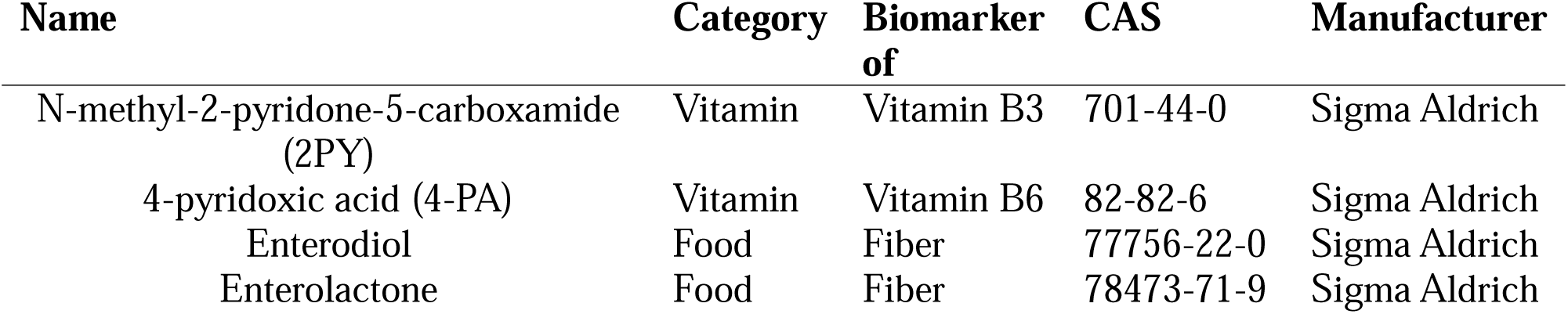
Biomarkers investigated in this study.

| **Name** | **Category** | **Biomarker**<br>**of** | **CAS** | **Manufacturer** |
| --- | --- | --- | --- | --- |
| N-methyl-2-pyridone-5-carboxamide<br>(2PY) | Vitamin | Vitamin B3 | 701-44-0 | Sigma Aldrich |
| 4-pyridoxic acid (4-PA) | Vitamin | Vitamin B6 | 82-82-6 | Sigma Aldrich |
| Enterodiol | Food | Fiber | 77756-22-0 | Sigma Aldrich |
| Enterolactone | Food | Fiber | 78473-71-9 | Sigma Aldrich |

**Table S2.** Concentration of biomarkers in the studied wastewater samples (N = 14).

| Location | Date | Biomarker (mg/kg) |  |  |  |
| --- | --- | --- | --- | --- | --- |
|  |  | 4-pyridoxic acid (4-PA) | Enterodiol | Enterolactone | N-methyl-2-pyridone-5-carboxamide (2PY) |
| MZUNI | 23-05-2025 | 1.5 | <0.3 | 8.6 | 81.8 |
| MZUNI | 24-05-2025 | 4.9 | <0.3 | 15.6 | 91.3 |
| MZUNI | 25-05-2025 | 4.7 | <0.3 | 4.8 | 94.9 |
| MZUNI | 26-05-2025 | 5.2 | <0.3 | 1.8 | 106.3 |
| MZUNI | 27-05-2025 | 5.2 | <0.3 | 2.7 | 121.7 |
| MZUNI | 28-05-2025 | 6 | <0.3 | 6 | 119.5 |
| MZUNI | 29-05-2025 | 6.5 | <0.3 | 5 | 89.9 |
| MUST | 16-06-2025 | 1.3 | <0.3 | 4.8 | 46.4 |
| MUST | 17-06-2025 | 13.4 | <0.3 | 7.3 | 39 |
| MUST | 18-06-2025 | 3.1 | <0.3 | 8 | 38.7 |
| MUST | 19-06-2025 | 2.2 | <0.3 | 8 | 34 |
| MUST | 20-06-2025 | 3.4 | <0.3 | 7.1 | 43.2 |
| MUST | 21-06-2025 | 5.7 | <0.3 | 7.2 | 56.2 |
| MUST | 22-06-2025 | 6.6 | <0.3 | 7.5 | 57.4 |
| LOD |  | 0.03 | 0.03 | 0.2 | 0.03 |
| LOQ |  | 0.1 | 0.3 | 0.5 | 0.1 |

## Text S1. Survey (English)

Demographics

1. What gender do you identify as? Male Female Non-binary /third gender Prefer not to say Other
2. What is your age range? Under 18 18 – 24 25 – 34 35 – 44 45 – 54 55 – 64 65 – 74 75 – 84 85 or older
3. What is the highest level of education you have completed? Primary school Secondary school Some University University degree Technical degree
4. What type of toilet do you use most often? Pit latrine Flush toilet No facility
5. What are your main sources of food; you/ your household? (Multiple choices)
  A. Only food purchased with own income
  B. Only food from your garden
  C. Food from your garden and others bought to supplement
  D. Supplement feeding with food from other community programs
  E. Other sources, please specify…………………………………………………
6. Please indicate with an X or a checkmark (√) <u>only the foods you ate during the past 24 hours</u>

7. How much is your budget for food in a normal month? ……………
8. If you had more money, list three foods you would buy/eat more?

|  |  |
| --- | --- |
| Group 1: All starchy staples diversity | Yes |
| Rice |  |
| Maize porridge |  |
| Pasta (Macaroni/spaghetti, etc.) |  |
| Maize on cob |  |
| Tubers (Plantains/green bananas, cassava, sweet potatoes, cocoyam/masimbi etc.) |  |
| Nsima from maize flour |  |
| Nsima from millet/sorghum flour |  |
| Nsima from cassava flour (kondowole) |  |
| Nsima from potato flour |  |
| Bread/Buns/Bread rolls |  |
| Scones/Biscuit/cake |  |
| Pastries (Doughnuts/mandazi etc.) |  |
| Rice porridge |  |
| Breakfast cereals (corn flakes etc.) |  |
| Oats/veetbix/instant porridge |  |
| Irish potatoes/chips |  |
| Sugar and other sweets |  |
| Candies /Jam and similar types |  |
| Soda (Fanta, coke, Sprite, etc) |  |
| Sweet potatoes |  |
| A beverage made from corn (Thobwa/chindongwa) |  |
| Other, please specify: |  |
| Group 2: Beans and peas | Yes |
| Dried beans |  |
| Dried pigeon peas (Nandolo/khobwe) |  |
| Soya |  |
| Chickpeas |  |
| Other, please specify: |  |
| Other, please specify: |  |
| Group 3: Nuts and Seeds | Yes |
| Peanut butter/peanuts |  |
| Other nuts (macadamia, etc.) |  |
| Pumpkin seeds |  |
| Sunflower seeds |  |
| Other, please specify: |  |
| Group 4: Dairy diversity | Yes |
| Evaporated/condensed milk |  |
| Cream or cheese |  |
| Butter (margarine) |  |
| Powdered milk |  |
| Milk, full cream, 2% or low fat (pasteurized or unpasteurized) |  |
| Custard |  |
| Sour cream (Chambiko) |  |
| Ice cream |  |
| Yoghurt |  |
| Soya pieces |  |
| Other, please specify: |  |
| Group 5: Flesh foods (meat, poultry, fish) diversity | Yes |
| Chicken |  |
| Beef (with/without bone), including steak, stew, ground) |  |
| Beef liver |  |
| Pork |  |
| Fish, fresh or frozen (tilapia, etc) |  |
| Tin Fish (sardines/tuna) |  |
| Sun dried fish (gelewa, bonya, kapenta, usipa etc.) |  |
| Smoked fish |  |
| Mutton (meat from sheep), including chops |  |
| Chicken livers/gizzards |  |
| Goat (meat) |  |
| Rabbits |  |
| Insects (ngumbi, locusts, crabs, mice, caterpillar<br>(mphalabungu/vilungulungu etc.) |  |
| Dried meat |  |
| Processed meat (polonies etc.) |  |
| Sausage |  |
| Bush meat |  |
| Other, please specify: |  |
| Group 6: Eggs diversity | Yes |
| Eggs (boiled, fried, poached, scrambled, omelette) |  |
| Group 7: Vegetables diversity | Yes |
| Rape |  |
| Spinach |  |
| Sweet potato leaves (kholowa/chibwaka) |  |
| Mango |  |
| Carrots |  |
| Orange flesh potatoes |  |
| Pumpkin |  |
| Papaya |  |
| Onions and other similar |  |
| Cabbage |  |
| Tomatoes |  |
| Green beans (fresh) |  |
| Peas (fresh – green) |  |
| Cauliflower |  |
| Pepper (red/green) |  |
| Hot pepper |  |
| Lettuce/mustard/rape/Chinese cabbage |  |
| Garlic |  |
| Fresh herbs (Ginger etc) |  |
| Cactus |  |
| Cucumber (Nkhaka) or horned melon (chipwete) |  |
| Amaranthus (bonongwe/belekete) |  |
| Blackjack (chisoso/kabata), |  |
| Bean leaves (khwanya) |  |
| Corchorus (Luni), |  |
| Pumpkin leaves (Nkhwani) |  |
| Cassava leaves (Chigwada) |  |
| Mushrooms |  |
| Limanda (Hibiscus) |  |
| Ocra (Thelele) |  |
| Other, please specify: |  |
| Group 8: Other fruits (and juices) diversity | Yes |
| Apple |  |
| Lemon |  |
| Orange |  |
| Tangerine |  |
| Banana |  |
| Pineapple |  |
| Avocado |  |
| Mulberries |  |
| Raspberry |  |
| Watermelon |  |
| Guava |  |
| Juices |  |
| Masau |  |
| Wild loquats (Masuku) |  |
| Other, please specify: |  |

……………………………..

…………………………….

………………………….…

Please select which answer is true for the last month? Remember members of your household include children.

|  |  |  |
| --- | --- | --- |
| 9 | In the last 4 weeks, did you worry that your household would not have enough food? | 0=No (Skip to Q2)<br>1=Yes |
|  | If yes, how often did this happen? | 1=Rarely (1-2 times)<br>2=Sometimes (3-10 times)<br>3=Often (more than 10 times) |
| 10 | In the last 4 weeks, were you or any household member not able to eat the kinds of foods you preferred because of a lack of resources? | 0=No (Skip to Q3)<br>1=Yes |
|  | If yes, how often did this happen? | 1=Rarely (1-2 times)<br>2=Sometimes (3-10 times)<br>3=Often (more than 10 times) |
| 11 | In the last 4 weeks, did you or any household member eat just a few kinds of food day after day because of lack of resources? | 0=No (Skip to Q4)<br>1=Yes |
|  | If yes, how often did this happen? | 1=Rarely (1-2times)<br>2=Sometimes (3-10times)<br>3=Often (more than 10 times) |
| 12 | In the last 4 weeks, did you or any household member eat some foods that you did not want to eat because of a lack of resources to obtain other types of food? | 0=No (Skip to Q5)<br>1=Yes |
|  | If yes, how often did this happen? | 1=Rarely (1-2times)<br>2=Sometimes (3-10 times)<br>3=Often (more than 10 times) |
| 13 | In the last 4 weeks, did you or any household member eat a smaller meal that you felt you needed more because there was not enough food? | 0=No (Skip to Q6)<br>1=Yes |
|  | If yes, how often did this happen? | 1=Rarely (1-2times)<br>2=Sometimes (3-10 times)<br>3=Often (more than 10 times) |
| 14 | In the last 4 weeks, did you or any household member eat fewer meals in a day because there was not enough food? | 0=No (Skip to Q7)<br>1=Yes |
|  | If yes, how often did this happen? | 1=Rarely (1-2times)<br>2=Sometimes (3-10times)<br>3=Often (more than 10 times) |
| 15 | In the last 4 weeks, was there ever no food at all in your household because there were no resources to get more? | 0=No (Skip to Q8)<br>1=Yes |
|  | If yes, how often did this happen? | 1=Rarely (1-2times)<br>2=Sometimes (3-10times)<br>3=Often (more than 10 times) |
| 16 | In the last 4 weeks, did you or any household member go to sleep at night hungry because there was not enough food? | 0=No (Skip to Q9)<br>1=Yes |
|  | If yes, how often did this happen? | 1=Rarely (1-2 times)<br>2=Sometimes (3-10 times)<br>3=Often (more than 10 times) |
| 17 | In the last 4 weeks, did you or any household member go a whole day without eating anything because there was not enough food? | 0=No (End)<br>1=Yes |
|  | If yes, how often did this happen? | 1=Rarely (1-2times)<br>2=Sometimes (3-10times)<br>3=Often (more than 10times) |

18. Common Symptoms of Deficiency. In the past 4 weeks, how often have you experienced the following symptoms?” Put a check in the column that applies with a Likert scale (e.g., Never – Rarely – Sometimes – Often – Always)

- Fatigue or low energy
- Irritability, depression, or mood swings
- Tingling or numbness in hands/feet (neuropathy)
- Muscle weakness
- Cracks or sores at corners of the mouth
- Glossitis (swollen, smooth tongue)
- Difficulty concentrating or memory issues
- Seizures
- Dermatitis (rash/ dark scaly skin on areas exposed to the sun)
- Vomiting and diarrhea
- Headache
- Apathy (lack of interest, enthusiasm, or concern)
- Disorientation (a state of mental confusion)
- Dizziness
- Weakened immune system (Frequent infection s/illness)
- Sore Throat
- Pale skin (in nail, beds, gums, inner lips, and inner eyelids appearing bluish gray)
- Constipation
- Skin lesions

## Text S2. Survey foods associated with biomarkers

Survey foods associated with the 2PY biomarker:

- Nsima from millet/ sourgum flour
- Bread/ Buns/ Bread rolls
- Eggs (boiled, fried, poached, scrambled, omelette)
- Peanut butter/ peanuts
- Chicken
- Beef (with/ without bone), including steak, stew, ground) Beef liver
- Pork
- Fish, fresh or frozen(tilapia, etc)
- Tin Fish (sardines/ tuna)
- Sun dried fish (gelewa, bonya, kapenta, usipa, etc.)
- Smoked fish
- Mutton (meat from sheep), including chops
- Chicken liver/ gizrads
- Goat
- Dried beans
- Dried Pigeon peas (Nandolo/ Khobwe)
- Soya
- Chickpeas
- Other nuts (macadamia, etc.)
- Pumpkin seeds
- Sunflower seeds

Survey foods associated with the 4-PA biomarker:

- Chicken
- Beef (with/ without bone), including steak, stew, ground) Beef liver
- Chicken liver/ gizrads
- Fish, fresh or frozen(tilapia, etc)
- Tin Fish (sardines/ tuna)
- Sun dried fish (gelewa, bonya, kapenta, usipa, etc.)
- Smoked fish
- Dried beans

Survey foods associated with the enterodiol or enterolactone biomarkers:

- Nsima from millet/ sourgum flour
- Dried beans
- Dried Pigeon peas (Nandolo/ Khobwe)
- Soya
- Chickpeas

